# Effects of the COVID-19 pandemic on TB outcomes in the United States: a Bayesian analysis

**DOI:** 10.1101/2024.10.17.24315683

**Authors:** Nicole A. Swartwood, Ted Cohen, Suzanne M. Marks, Andrew N. Hill, Garrett R. Beeler Asay, Julie Self, Pei-Jean I. Feng, C. Robert Horsburgh, Joshua A. Salomon, Nicolas A. Menzies

## Abstract

**Background:** Tuberculosis (TB) cases and deaths in the United States fluctuated substantially during the COVID-19 pandemic. We analyzed multiple data sources to understand the factors contributing to these changes and estimated future TB trends.

**Methods:** We identified four mechanisms potentially contributing to observed TB trends during 2020– 2023: immigration, respiratory contact rates, rates of accurate diagnosis and treatment initiation, and mortality rates for persons with TB disease. We employed a Bayesian approach to synthesize evidence on how these mechanisms changed during the pandemic and how they might have combined to produce observed 2020–2023 TB data, using a transmission-dynamic model to link mechanisms to TB outcomes. We also simulated a no-pandemic counterfactual scenario that assumed mechanisms followed pre-pandemic trends. We estimated TB outcomes associated with the pandemic until 2035 to capture lagged effects. We evaluated additional scenarios to estimate the individual effect of each mechanism.

**Results:** Over the 2020–2035 study period, we estimate an additional 2,784 (95% uncertainty interval: 2,164–3,461) TB cases and 1,138 (1,076–1,201) TB deaths in the United States associated with changes occurring during the COVID-19 pandemic. The four mechanisms had offsetting effects – decreases in TB diagnosis rates and increases in TB mortality rates led to more TB deaths, while reductions in contact rates reduced TB deaths. Immigration changes initially reduced TB deaths, but increased deaths over time.

**Discussion:** While the direct impacts of the COVID-19 pandemic occurred between 2020–2023, these changes may continue to influence TB incidence and mortality in future years.

**Summary:** The COVID-19 pandemic disrupted steady declines in U.S. tuberculosis rates. Despite sharp decreases in TB cases over 2020–2021, we projected there would be 2,784 additional TB cases and 1,138 additional TB deaths during 2020–2035 associated with pandemic-related changes.

## Background

Preliminary tuberculosis (TB) surveillance data suggest that United States (U.S.) TB cases increased 16% in 2023 compared to 2022 [1]. This rise occurred three years after a 19% reduction in TB cases in 2020 compared to 2019 [2]. These fluctuations represent notable departures from the slow decline in TB cases observed over the prior eight years (2011–2019), during which annual percentage changes ranged from -5.4% to 1.7% [3]. While TB deaths declined by 2% between 2018 and 2019, TB mortality increased sharply following 2019, with the number of persons dying with TB 17% and 21% higher in 2020 and 2021, respectively, compared to 2019 [4]. This variation in TB cases and deaths could have resulted from changes related to the COVID-19 pandemic.

The COVID-19 pandemic disrupted U.S. health services [5, 6], including routine and preventive TB care [7, 8]. Resulting diagnostic delays and treatment interruptions might have increased the duration of infectiousness for TB disease and delayed introduction of life-saving therapy, prolonging morbidity and leading to elevated mortality rates compared to the pre-pandemic period [9, 10]. COVID-19 co-infection might have also led to worse outcomes, including increased mortality, for persons with TB [11, 12].

The COVID-19 pandemic might also have affected TB through changes in immigration. Since 2002 the majority of TB cases in the United States have occurred among non-U.S.–born persons [3]. In 2022, non-U.S–born persons accounted for 72% of total reported TB cases, and 31% of these non-U.S–born cases occurred within four years since entry [3]. As such, TB rates in the United States are sensitive to the number and origin of migrants entering each year [13]. The initial federal response to COVID-19 included provisions that limited immigration into the United States [14]. These efforts led to fewer immigrants between March 2020 and February 2021, followed by a rebound in March 2021 through September 2022 once pandemic measures were relaxed [15]. These immigration changes might also affect long-term TB incidence as 51% of TB cases among non-U.S.–born persons occur in persons who have been in the United States over ten years [3].

Changes in TB transmission during the pandemic might have affected TB epidemiologic trends. Both SARS-CoV-2 and *Mycobacterium tuberculosis* (*Mtb*) are spread through respiratory transmission [16]. For this reason, the non-pharmaceutical interventions designed to limit SARS-CoV-2 transmission—such as stay at home orders, physical distancing and face mask use— might have lowered respiratory contact rates, and reduced TB transmission [17, 18].

As these factors were changing simultaneously over the pandemic period, it is difficult to estimate their individual impacts on TB dynamics in the United States based on observational data alone. However, understanding their potential effect on U.S. TB (collectively and individually) is important for predicting future trends and preparing for potential future pandemics. Mathematical models have proven a useful tool for investigating these types of questions, by allowing multiple different evidence sources to be synthesized in an internally consistent framework, constraining estimated outcomes to align with existing epidemiological evidence, and allowing future trends to be forecast under multiple counterfactual scenarios [19, 20].

In this study, we used a transmission dynamic model to investigate the potential impact of COVID-19-associated changes on U.S. TB outcomes. We focused on four potential mechanisms through which COVID-19 might have impacted TB dynamics—changes in immigration, respiratory contact rates, rates of accurate TB diagnosis and treatment initiation (access to care), and mortality rates among persons with TB—and synthesized available data describing these changes over 2020-2023. We estimated the individual and collective impact of these mechanisms on *Mtb* infections, TB cases, and TB deaths, both during the pandemic period (2020–2023) and over future years (2024–2035), compared to a no-pandemic counterfactual. We used these estimates to describe the potential long-term impacts of COVID-19 pandemic on TB in the United States.

## Methods

### Reported TB data

We collated annual data on four outcomes describing changes in U.S. TB during 2020–2023: total TB cases, the percentage of TB cases occurring in non-U.S.–born persons, the percentage of non-U.S.–born TB cases occurring in new arrivals (entered the United States <2 years before notification year), and TB deaths. We extracted data on TB cases for 2020-2022 from the National TB Surveillance System (NTSS), which contains TB notifications reported by the 50 U.S. states and District of Columbia [3]. We retained variables describing the year the case was counted, origin of birth (U.S.-born or non-U.S.–born), and length of time in the United States. NTSS defines U.S.-born persons as individuals entitled to U.S. citizenship at birth. TB case data for 2023 were extracted from preliminary reporting by the Centers for Disease Control and Prevention (CDC) [1]. Reported TB deaths during 2020–2023 were extracted from CDC’s Multiple Cause of Death database [4]; all deaths with TB listed as a cause of death (underlying or non-underlying) on the death certificate were used. These are referred to as “TB deaths.”

### Data on epidemiological mechanisms affected by the COVID-19 pandemic

We collated data on each of the four epidemiological mechanisms potentially affected by the COVID-19 pandemic. Data describing changes in immigration during March 2020–September 2022 were extracted from Department of Homeland Security reports [15]. We used data on numbers of Legal Permanent Resident New Arrival admissions and assumed rates of entry for other migrant categories changed proportionally to these values. Estimated changes in respiratory contact rates during the pandemic were informed by interpersonal contact rates estimated by a previous study [21]. To allow for changes during 2020–2023, we assumed that the overall reduction in contact rates was proportional to the Oxford Stringency Index, a time-varying composite measure of COVID-19 mitigation measures [22]. Data on reductions in access to care were collected from CDC’s Research and Development Survey [23], and we assumed changes in the rate of TB diagnosis and treatment (for persons with undiagnosed disease) were proportional to these values.

### Mathematical model and parameter calibration

We adapted a published mathematical model of TB [24], which simulates *Mtb* transmission and TB progression within the United States as a function of demographic trends, risk factor distributions, immigration patterns, and the provision of TB prevention and treatment services, with a monthly timestep. The model was fit to data on U.S. demography, reported TB epidemiological data, natural history evidence, and health services data spanning 1950–2019.

We used this model to synthesize evidence describing changes in epidemiological mechanisms and reported TB outcomes (cases and deaths) during 2020–2023. We incorporated this evidence a Bayesian calibration approach, in which prior distributions were created to represent time trends in each of the four epidemiological mechanisms over 2020–2023, and calibration targets created from the reported TB data.

We parameterized changes in each epidemiological mechanism as a step function, with steps every six months between March 2020 and March 2023 (approximating the expiration of the federal Public Health Emergency for COVID-19 [25]), allowing six steps per mechanism (Supplementary Table 1). We assumed that each mechanism would return linearly to projected pre-pandemic levels between March and December 2023.

We defined log-likelihood functions comparing model estimates to reported data for annual TB cases, TB deaths, percentage of TB cases occurring in non-U.S.–born persons, and the percentage of non-U.S.–born TB cases among new arrivals. We used incremental mixture importance sampling to sample the posterior distribution for the step function parameters of each epidemiological mechanism [26]. This procedure yielded 1,000 parameter sets consistent with the reported TB data while representing the joint uncertainty in the effects of the four epidemiological mechanisms.

### Analytic scenarios

We used the calibrated model to create a ‘fitted pandemic scenario’, representing actual TB outcomes during the COVID-19 pandemic, and used this scenario to project annual TB outcomes (incident *Mtb* infections, reported TB cases, and TB deaths) through 2035. We also calculated outcomes for a counterfactual ‘no-pandemic’ scenario, which assumed that the four epidemiological mechanisms followed pre-pandemic trends unaffected by the pandemic. For immigration, we assumed no net increase in immigration volume during the pandemic period, i.e., cumulative immigration in the no-pandemic counterfactual is equal to that under the fitted pandemic scenario. We ran additional scenario analyses exploring the impact of a ±25% change in immigration during 2020–2023. For the remaining mechanisms, we assumed the pre-pandemic trends were a continuation of observed trends through 2019.

We calculated the difference in incident *Mtb* infections, reported TB cases, and TB deaths between the fitted pandemic scenario and the no-pandemic scenario to quantify the impact of the COVID-19 pandemic on U.S. TB outcomes (representing the combined effect of the four epidemiological mechanisms) between 2020 and 2035.

We created additional scenarios to explore the contribution of each individual epidemiological mechanism to the overall pandemic effect. These scenarios were created by modifying the fitted pandemic scenario such that a single epidemiological mechanism was assumed to follow its ‘no-pandemic’ trend, while the other three followed the fitted pandemic scenario. We calculated the difference in TB outcomes under each alternative scenario and the fitted pandemic scenario during 2020–2035 to estimate the TB cases and TB deaths associated with each epidemiological mechanism. As the mathematical model accounted for the non-linear interaction of epidemiological mechanisms, the sum of these four effects was not expected to sum to the total pandemic effect.

### Statistical analysis

We simulated outcomes for each of 1000 parameter samples. We calculated point estimates as the median of these simulated results and reported equal-tailed 95% uncertainty intervals as the 2.5^th^ and 97.5^th^ percentiles of the distribution of results. All analyses were conducted in R (v. 4.2.1) using the Rcpp and MITUS R packages [27–29].

## Results

### Fitted pandemic scenario

Figure 1 shows the monthly modeled trend in the mechanism parameters compared with the reported data used to inform the model. Supplementary Figure 1 compares results of the fitted model to reported TB data over the 2020–2023 period.

**Figure 1:**
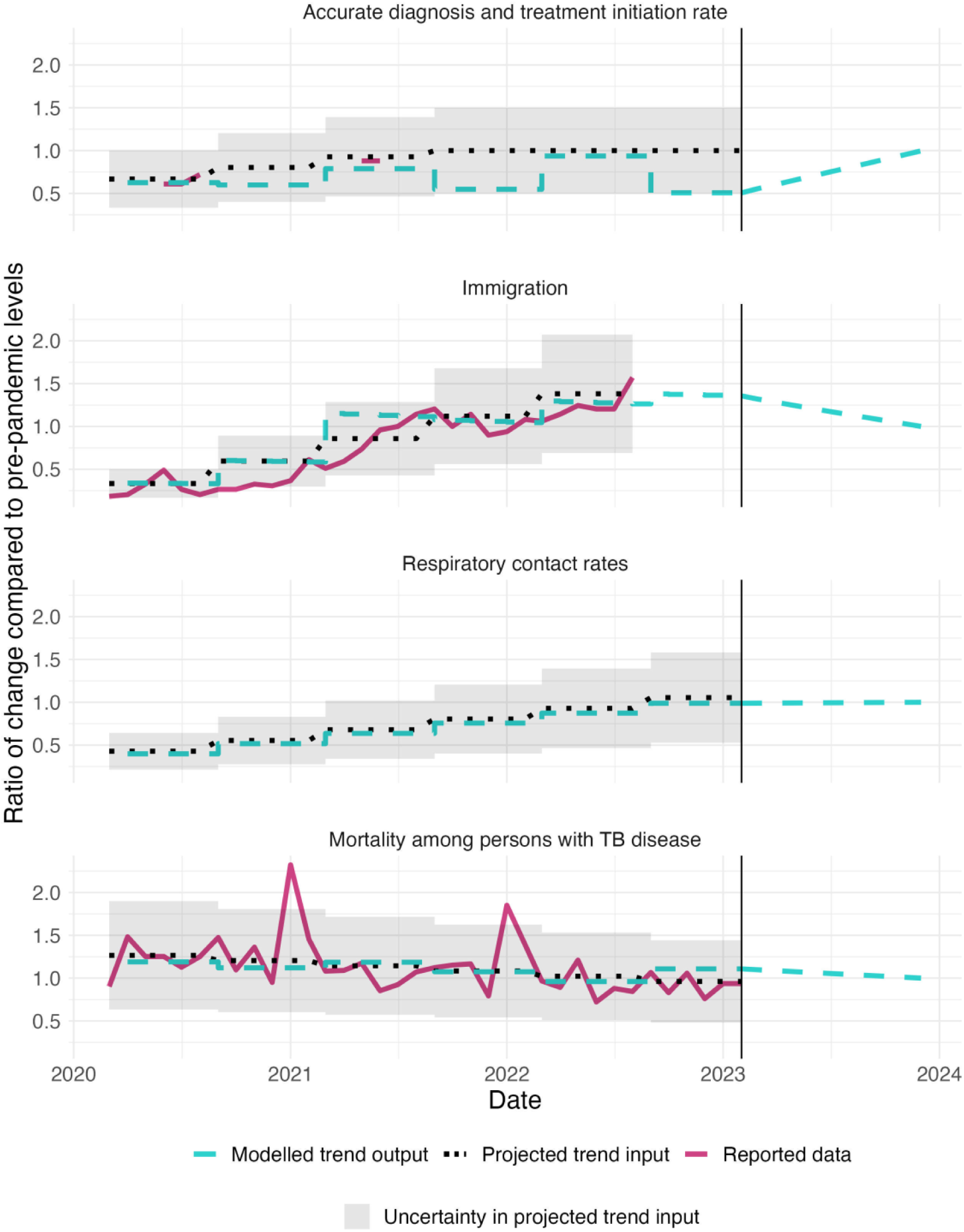
Comparison of reported trends in COVID-19 adjustment mechanisms to projected trend inputs and modelled trend outputs for each of the four examined mechanisms during 2020–2023. Note: 1.0 represents equality with pre-pandemic projected levels. As respiratory contact trend is a synthesis of multiple data sources, no reported data is shown for that mechanism. Y-axis is different across parameters.

### Future TB outcomes

Figure 2 shows modelled TB outcomes projected to 2035 under the fitted pandemic scenario and no-pandemic counterfactual. During 2020–2035, cumulative cases were estimated to be 132,417 (95% uncertainty interval: 128,670–137,403) under the no-pandemic counterfactual and 135,187 (131,189–140,493) under the fitted pandemic scenario, a difference of 2.1% (1.9–2.3%), or 2,784 (2,164–3,461) additional cases. A majority (74.7% [74.0–75.4%]) of the additional TB cases associated with the pandemic were estimated to occur among non-U.S.–born persons, consistent with pre-pandemic trends.

**Figure 2:**
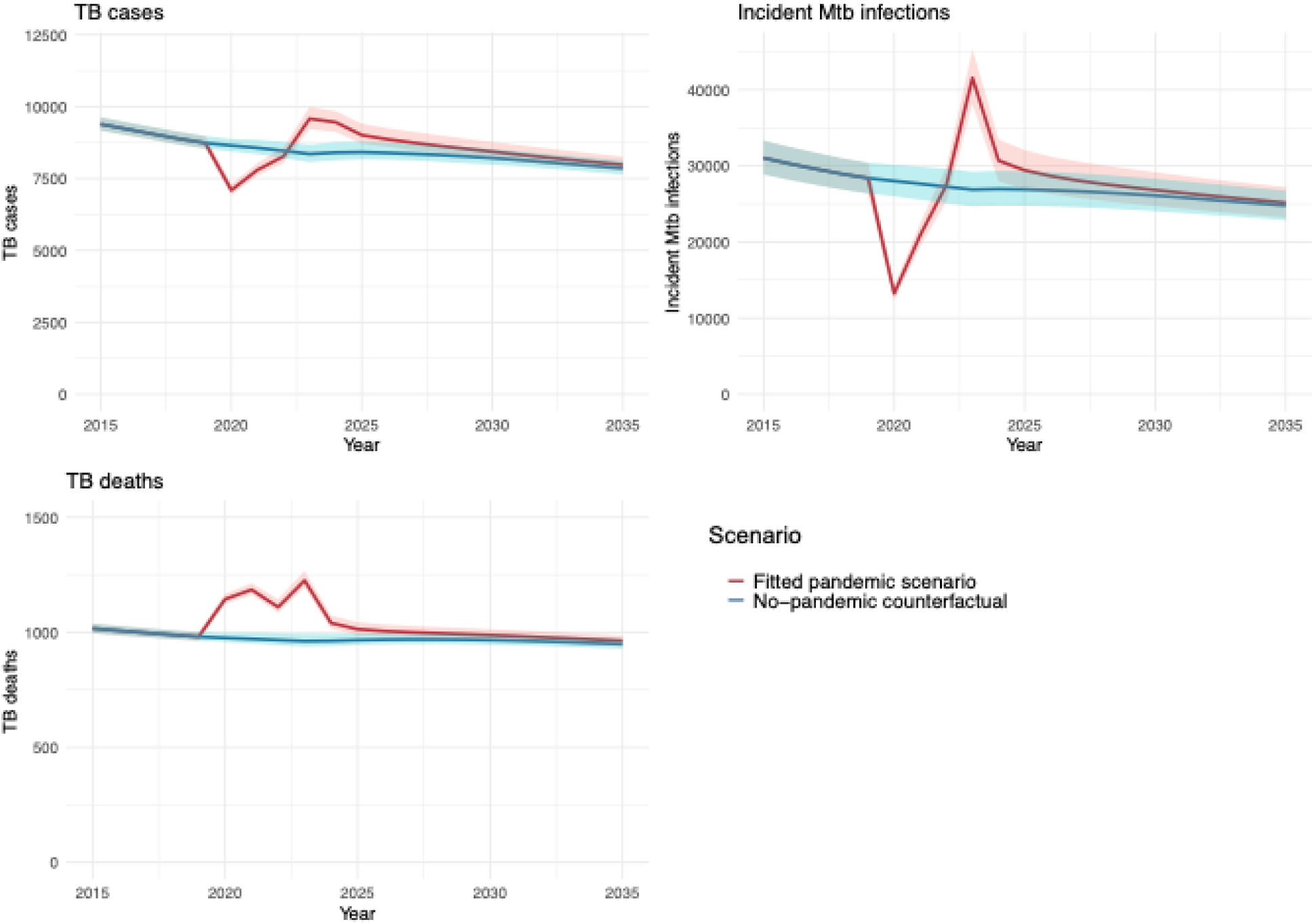
Model estimated annual TB outcomes under the fitted pandemic scenario and the no-pandemic counterfactual during 2015–2035.* * Solid lines represent point estimates. Shaded regions represent 95% uncertainty intervals.

Table 1 presents the estimated annual differences in TB outcomes between the fitted pandemic scenario and the no-pandemic counterfactual for selected years. Under the fitted pandemic scenario, annual TB cases were lower over 2020–2022 compared to the counterfactual, resulting in a cumulative 2,500 (2,333–2,682) fewer TB cases during these years. Over 2023–2035, annual TB cases were higher under the fitted pandemic scenario compared to the no-pandemic scenario (5,284 ([4,845–5,794]) additional TB cases).

**Table 1:** Net changes in TB outcomes associated with the COVID-19 pandemic in selected calendar years, stratified by nativity. Numbers in parentheses represent 95% uncertainty intervals.

|  | Calendar year |  |  |  |  |  |
| --- | --- | --- | --- | --- | --- | --- |
|  | 2020 | 2021 | 2022 | 2023 | 2025 | 2035 |
| ***TB cases*** |  |  |  |  |  |  |
| *Total* | -1,551<br>(-1,622, -1,493) | -760<br>(-832, -691) | -189<br>(-227, -148) | 1,228<br>(1,151, 1,332) | 593<br>(542, 652) | 122<br>(106, -139) |
| *U.S.-born* | -410<br>(-427, -392) | -251<br>(-276, -226) | -196<br>(-215, -174) | 228<br>(205, 252) | 282<br>(252, 311) | 28<br>(23, 31) |
| *non-U.S.-born* | -1,141<br>(-1,244, -1,077) | -509<br>(-573, -453) | 7<br>(-44, 52) | 1,000<br>(935, 1,093) | 311<br>(282, 349) | 94<br>(83, -107) |
| ***Incident Mtb infections*** |  |  |  |  |  |  |
| *Total* | -14,769<br>(-15,957, -13,672) | -6,733<br>(-7,365, -6,105) | 228<br>(59, 421) | 14,683<br>(13,437, 16,125) | 2,513<br>(2,191, 2,917) | 368<br>(307, 448) |
| *U.S.-born* | -9,075<br>(-9,729, -8,492) | -3,708<br>(-4,028, -3,421) | 652<br>(561, 763) | 10,435<br>(9,650, 11,373) | 1,173<br>(1,041, 1,354) | 161<br>(127, 199) |
| *non-U.S.-born* | -5,694<br>(-6,228, -5,108) | -3,025<br>(-3,338, -2,685) | -424<br>(-502, -342) | 4,248<br>(3,787, 4,752) | 1,340<br>(1,150, 1,563) | 207<br>(180, 249) |
| ***TB deaths*** |  |  |  |  |  |  |
| *Total* | 169<br>(163, 174) | 215<br>(207, 222) | 143<br>(136, 149) | 263<br>(252, 277) | 47<br>(43, 52) | 14<br>(13, 15) |
| *U.S.-born* | 43<br>(41, 45) | 49<br>(46, 52) | 25<br>(23, 27) | 52<br>(50, 55) | 17<br>(15, 19) | 2<br>(2, 2) |
| *non-U.S.-born* | 126<br>(122, 129) | 166<br>(161, 170) | 118<br>(113, 122) | 211<br>(202, 222) | 30<br>(28, 33) | 12<br>(11, 13) |

Note: a negative number represents lower TB outcomes estimated for the fitted pandemic scenario compared to the no-pandemic counterfactual.

During 2020–2035, 15,428 (15,095–15,844) cumulative TB deaths were estimated under the no-pandemic counterfactual and 16,566 (16,203–17,016) deaths under the fitted pandemic scenario. The estimated 1,138 (1,076–1,201) additional deaths represented a 7.4% (7.2–7.5%) cumulative increase compared with the no-pandemic counterfactual. Under both the fitted pandemic scenario and no-pandemic counterfactual the majority of deaths with TB were among persons who were non-U.S.–born, comprising 80.3% (79.7–80.9%) and 80.5% (80.0–81.2%), respectively. Under the fitted pandemic scenario, estimated annual TB deaths were elevated across the full study period, compared with the no-pandemic counterfactual.

During 2020–2035, an estimated 430,783 (396,390–467,892) incident *Mtb* infections occurred under the fitted pandemic scenario and 422,752 (389,523–458,475) under the no-pandemic counterfactual, representing a 1.9% (1.6–2.3%) increase.

### Impact of individual epidemiological mechanisms

Figure 3A shows the total changes in TB cases and deaths attributable to the changes in each epidemiological mechanism during the pandemic, estimated over the 2020-2035 study period.

**Figure 3:**
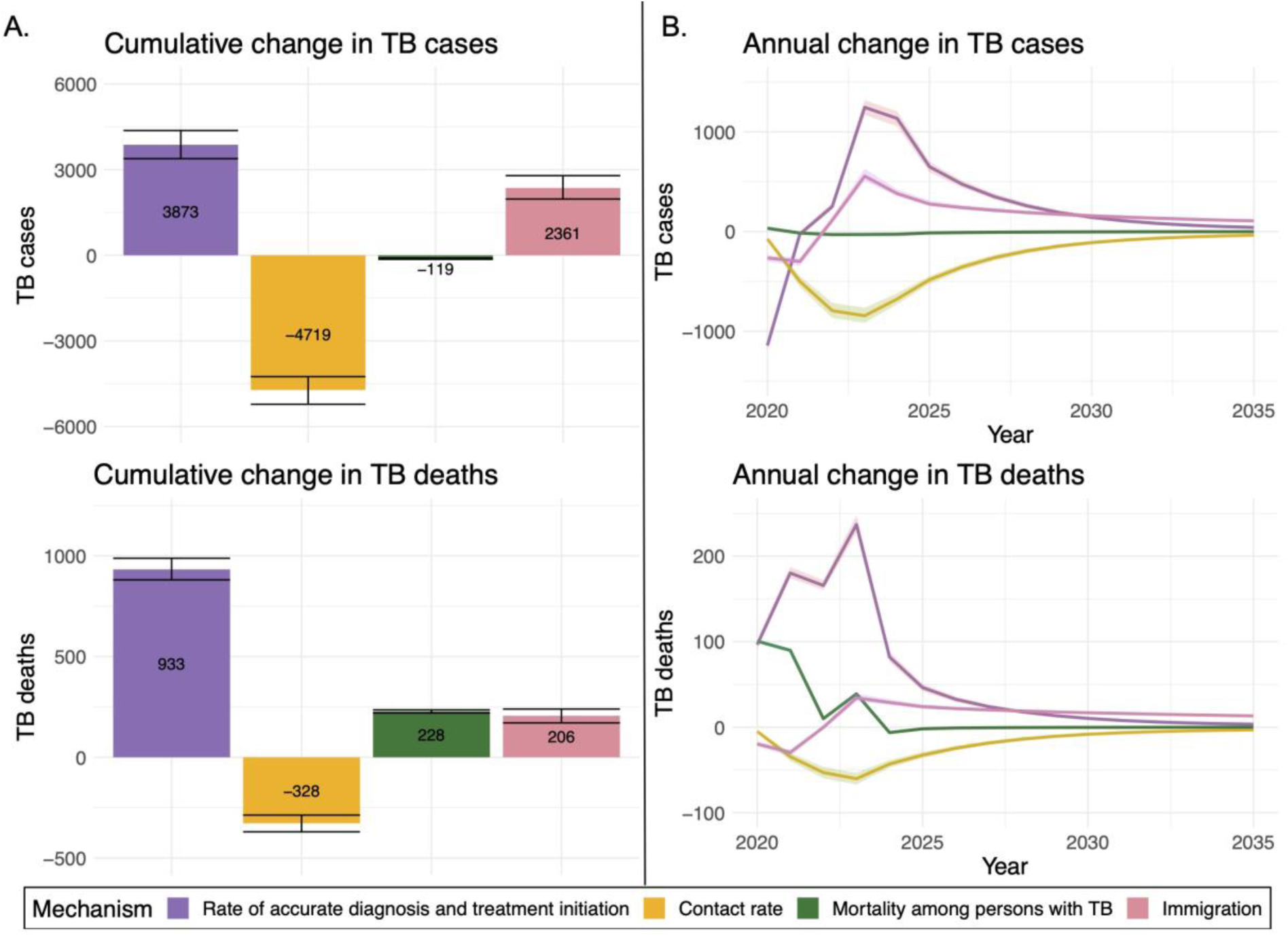
Estimated impact of each examined COVID-19 related mechanism on TB outcomes during 2020–2035. A) Cumulative difference in TB outcomes associated with each mechanism B) Annual difference in TB outcomes associated with each mechanism. Note: Error bars and shaded area represent 95% uncertainty intervals.

Figure 3B depicts time trends in TB cases and deaths associated with each mechanism. For several of these mechanisms we estimated most cumulative mechanism-associated impacts on TB outcomes would occur during 2024–2035, after parameters returned to pre-COVID levels. During 2024–2035, an estimated 3,545 (3,230–3,869) and 2,255 (2,029–2,520) additional TB cases were associated with pandemic-related changes in rates of accurate diagnosis and treatment initiation and immigration, respectively. In contrast, pandemic-related changes in respiratory contact rates were estimated to produce 2,510 (2,260–2,797) fewer TB cases during 2024–2035. Change in TB mortality had its effect on TB cases concentrated during 2020–2023. The impact of pandemic related changes in mechanisms on TB deaths followed similar trends to TB cases.

Supplementary Figure 2 shows the differences in TB outcomes under a sensitivity analysis that varied no-pandemic counterfactual immigration. A 25% reduction in counterfactual (no-pandemic) immigration during 2020–2023 resulted in 3,639 (3,240–4,256) additional TB cases, 344 (305–408) additional TB deaths and 11,503 (10,050 –13,792) under the fitted pandemic scenario over 2020–2035. A 25% increase in counterfactual immigration during 2020–2023 resulted in 4,469 (4,455 –4635) fewer TB cases, 422 (415–434) fewer TB deaths and 13,295 (11,503–15,107) under the fitted pandemic scenario over 2020 –2035.

## Discussion

The COVID-19 pandemic caused widespread disruption within the United States during 2020– 2023. In this analysis, we examined pandemic impacts on TB outcomes—as mediated through changes in immigration, respiratory contact rates, access to TB care, and mortality among persons with TB disease—during the 2020-2023 period and modelled long-term effects of the pandemic on TB epidemiology in the United States. During 2020–2035, we estimated 2,770 additional TB cases and 1,138 additional TB deaths due to pandemic-associated changes in TB epidemiology. These overall changes were produced by an initial sharp decrease in TB cases during 2020-2022, followed by an increase in TB cases through 2035. The estimated increase in TB cases might be due, at least in part, to a “catch-up” in the diagnosis of TB cases not diagnosed due to lack of access to TB care or changes in the timing of immigration [1]. Estimated additional deaths due to TB might reflect the elevated frequency of death among those with COVID-19 co-infection and increased disease severity at the time of TB diagnosis, possibly associated with delayed diagnoses [7, 12].

The United States provides a valuable case study of the impact of COVID-19 on TB, in a setting where TB trends are largely driven by reactivation of LTBI among non-U.S.–born persons [3]. Our analysis suggests that the COVID-19 pandemic influenced TB among U.S.-born and non-U.S.–born persons through different mechanisms, demonstrating a mathematical relationship between immigration rates and TB incidence rates among non-U.S.–born persons. The sharp decrease in immigration in 2020 was associated with fewer TB cases among non-U.S.–born persons during 2020–2022. TB cases among non-U.S.–born persons then subsequently increased as TB services returned to pre-pandemic levels and reported immigration rebounded. This positive relationship between migration volume and non-U.S.–born TB rates reflects the disproportionate contribution of recent migrants to U.S. TB case totals, with rates of TB diagnosis for migrants in the years immediately following U.S. entry substantially higher than for U.S.-born persons and long-term residents [30]. In 2023, modeled TB cases totals among non-U.S.–born persons surpassed their modelled no-pandemic counterfactual projection, reflecting increased immigration during 2021–2022 [15].

A number of studies that have investigated the impact of COVID-19 on TB in high incidence settings, but few studies have been published for the United States [7, 10, 12, 31, 32]. An analysis of the 2020–2021 Global Burden of Disease (GBD) estimates reported excess TB deaths among persons under 65 years of age, resulting in an observed to expected deaths ratio of 1.09 (0.983 to 1.20) across all ages [33]. Our model estimates suggest a ratio of 1.20, which falls at the high end of the reported range. This elevated estimate might be due to the exclusion of persons living with HIV in the GBD estimate, a population which has been shown to have increased mortality among persons with TB [12]. Furthermore, we can compare our fitted parameter estimates to empirical metrics. For instance, our estimates of reductions in accurate diagnosis and treatment initiation are consistent with an analysis which found a 37% decrease in sputum smear and culture samples collected during 2020 in two large Californian hospitals [34]. An analysis of emergency department claims among Medicare beneficiaries also found a decrease in TB claims during 2020 [32]. Additionally, the number of reported persons who were identified as contacts of a TB case declined in 2020 [35].

This analysis has several limitations. First, our estimates are sensitive to the assumptions and data used for model fitting, the latter of which were not available for all mechanisms over the total 2020–2023 period. Data used to inform trends in the rates of diagnosis and treatment initiation were only available at a few time points. In contrast, our study benefited from the high quality of U.S. TB surveillance data, which allowed us to calibrate the model to multiple epidemiological metrics and time-points. Secondly, our model did not account for potential changes in TB burden among persons immigrating to the United States. Disruptions in TB care in sender countries might result in increased LTBI incidence among persons who immigrate to the United States [36]. Similarly, the pandemic might have produced shifts in the types of individuals migrating to the United States [37], which could lead to changes in TB rates after U.S. entry. Third, our study chose to explore each mechanism at a national level. However, both TB outcomes and COVID-19 impacts vary substantially across sub-national geographies [38, 39]. Further research examining the sub-national impact of the pandemic on TB outcomes could better inform future public health responses.

This study used mathematical modeling to explore the impact of COVID-19 pandemic on TB in the United States, finding increases in both TB cases and TB deaths during 2020–2035. While the direct impacts of the COVID-19 pandemic occurred between 2020–2023, our analysis suggests these changes might continue to influence TB incidence and mortality in future years.

## Data Availability

Apart from National TB Surveillance System data, all data used in this analysis represent deidentified publicly available datasets. National TB Surveillance System data contain information abstracted from the national tuberculosis case report form called the Report of Verified Case of Tuberculosis (RVCT) (OMB No. 0920-0728). These data have been reported voluntarily to CDC by state and local health departments and are protected under the Assurance of Confidentiality (Sections 306 and 308(d) of the Public Health Service Act, 42 U.S.C. 242k and 242m(d)), which prevents disclosure of any information that could be used to directly or indirectly identify patients. For more information, see the CDC/ATSDR Policy on Releasing and Sharing Data (at https://stacks.cdc.gov/view/cdc/7563). A limited dataset is available at http://wonder.cdc.gov/tb.html. Researchers seeking additional National TB Surveillance System data may request access through the National Center for Health Statistics Research Data Centers (https://www.cdc.gov/rdc/b1datatype/tuberculosis.htm).

## Acknowledgements

We thank Stephanie Su and Teresa Puente for assistance in study coordination.

## Author Contributions

N.A.S., N.A.M., and S.M.M. conceived the project. N.A.M. and J.A.S. acquired funding. N.A.S. wrote the model code, drafted the original manuscript, and visualized the results. N.A.S. curated the data and executed the analysis. All authors contributed to the development of the methodology, and reviewed and edited the original manuscript.

## Disclaimer

The findings, conclusions, and views expressed are those of the authors and do not necessarily represent the official position of the Centers for Disease Control and Prevention (CDC).

## Data sharing statement

Apart from National TB Surveillance System data, all data used in this analysis represent deidentified publicly available datasets. National TB Surveillance System data contain information abstracted from the national tuberculosis case report form called the Report of Verified Case of Tuberculosis (RVCT) (OMB No. 0920-0728). These data have been reported voluntarily to CDC by state and local health departments and are protected under the Assurance of Confidentiality (Sections 306 and 308(d) of the Public Health Service Act, 42 U.S.C. 242k and 242m(d)), which prevents disclosure of any information that could be used to directly or indirectly identify patients. For more information, see the CDC/ATSDR Policy on Releasing and Sharing Data (at https://stacks.cdc.gov/view/cdc/7563). A limited dataset is available at http://wonder.cdc.gov/tb.html. Researchers seeking additional National TB Surveillance System data may request access through the National Center for Health Statistics’ Research Data Centers (https://www.cdc.gov/rdc/b1datatype/tuberculosis.htm). N.A.S. had full access to all the data in the study and takes responsibility for the integrity of the data and the accuracy of the data analysis.

