## Supplementary for "Effects of the COVID-19 pandemic on TB outcomes in the United States: a Bayesian analysis"

**Contents**

I. Table S1: Calibrated baseline and COVID-19 mechanism adjustment parameters used in the transmission model.
II. Figure S1: Comparison of calibration targets and model estimates for the fitted pandemic scenario and no-pandemic counterfactual during 2019–2023. Shaded areas represent 95% uncertainty intervals.
III. Figure S2: Model estimated annual difference in TB outcomes under the fitted pandemic scenario compared with each counterfactual immigration scenario during 2015–2035.

| **Parameter name** | **Parameter mean and 95% interval** | **Prior distribution informed by reported data** | **Sources** |
| --- | --- | --- | --- |
| *Accurate diagnosis and treatment initiation rates* | | | |
| Baseline value | 0.30 | | Model calibrated |
| Time varying ratio of change  *March – August 2020* | 0.67 (0.38, 1.04) | Gamma(15.19, 22.76) | [1] |
| *September 2020 – February 2021* | 0.80 (0.45, 1.25) | Gamma(15.19, 18.91) |  |
| *March – August 2021* | 0.92 (0.52, 1.45) | Gamma(15.19, 16.37) |  |
| *September 2021 – February 2022* | 1.00 (0.56, 1.56) | Gamma(15.19, 15.19) |  |
| *March – August 2022* | 1.00 (0.56, 1.56) | Gamma(15.19, 15.19) |  |
| *September 2022 – February 2023* | 1.00 (0.56, 1.56) | Gamma(15.19, 15.19) |  |
| *Respiratory contact rates* | | | |
| Baseline value | Varies by nativity and risk;  ranges from 0.00043 (low risk, US-born)  to 0.036 (high risk non-U.S.–born) | | Model calibrated |
| Time varying ratio of change  *March – August 2020* | 0.42 (0.24, 0.67) | Gamma(15.19, 35.43) | [2, 3] |
| *September 2020 – February 2021* | 0.55 (0.31, 0.87) | Gamma(15.19, 27.42) |  |
| *March – August 2021* | 0.68 (0.38, 1.06) | Gamma(15.19, 22.37) |  |
| *September 2021 – February 2022* | 0.80 (0.45, 1.25) | Gamma(15.19, 18.89) |  |
| *March – August 2022* | 0.93 (0.52, 1.45) | Gamma(15.19, 16.34) |  |
| *September 2022 – February 2023* | 1.05 (0.59, 1.64) | Gamma(15.19, 14.40) |  |
| *TB mortality rate* | | | |
| Baseline value | Varies with age; ranges from  0.0013 (0-4 years) to 0.17 (95+ years) | | Model calibrated |
| Time varying ratio of change  *March – August 2020* | 1.26 (0.71, 2.00) | Gamma(15.19, 12.00) | [4, 5] |
| *September 2020 – February 2021* | 1.21 (0.68, 1.88) | Gamma(15.19, 12.61) |  |
| *March – August 2021* | 1.14 (0.64, 1.79) | Gamma(15.19, 13.28) |  |
| *September 2021 – February 2022* | 1.08 (0.61, 1.69) | Gamma(15.19, 14.03) |  |
| *March – August 2022* | 1.02 (0.57, 1.59) | Gamma(15.19, 18.86) |  |
| *September 2022 – February 2023* | 0.96 (0.54, 1.50) | Gamma(15.19, 15.81) |  |
| *Immigration volume* | | | |
| Baseline value | 132,705 | | Model calibrated |
| Time varying ratio of change  *March – August 2020* | 0.33 (0.19, 0.52) | Gamma(15.19, 45.57) | [6] |
| *September 2020 – February 2021* | 0.60 (0.33, 0.93) | Gamma(15.19, 25.51) |  |
| *March – August 2021* | 0.85 (0.28, 1.34) | Gamma(15.19, 17.72) |  |
| *September 2021 – February 2022* | 1.12 (0.63, 1.75) | Gamma(15.19, 13.57) |  |
| *March – August 2022* | 1.38 (0.78, 2.15) | Gamma(15.19, 11.00) |  |
| *September 2022 – February 2023* | 1.64 (0.92, 2.57) | Gamma(15.19, 9.24) |  |

**Table S1: Calibrated baseline and COVID-19 mechanism adjustment parameters used in the transmission model.**

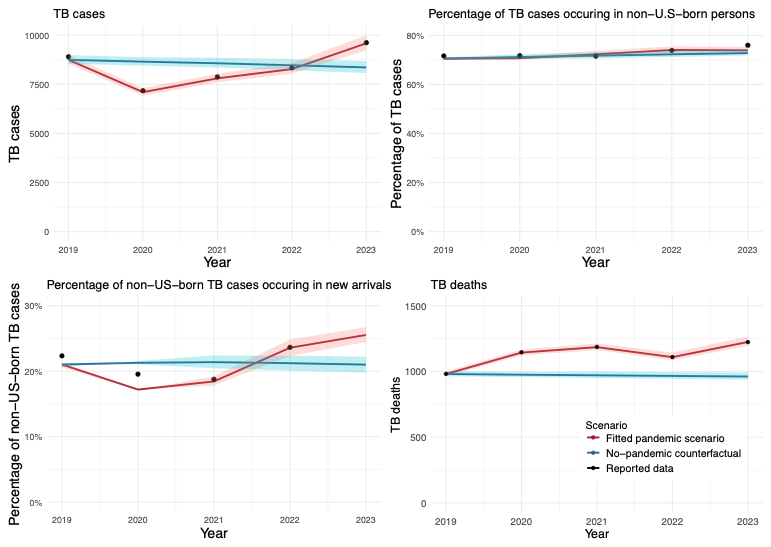

**Figure S1: Comparison of calibration targets and model estimates for the fitted pandemic scenario and no-pandemic counterfactual during 2019–2023. Shaded areas represent 95% uncertainty intervals.**

Note: New arrivals defined as non-U.S.–born persons in the United States less than or equal to 2 years.

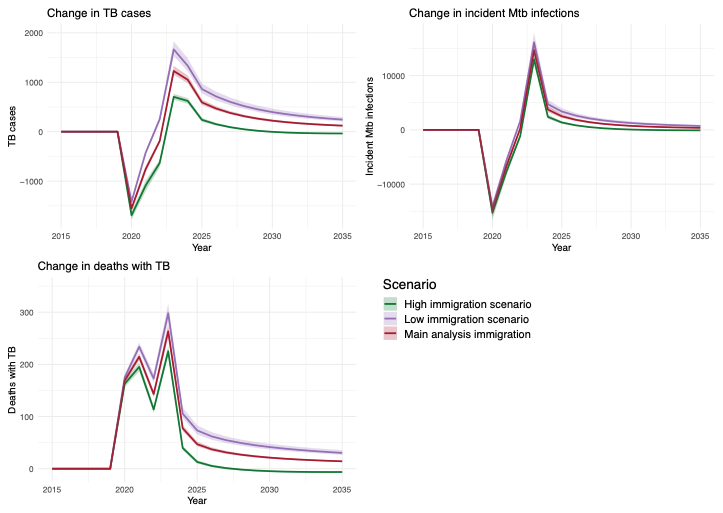

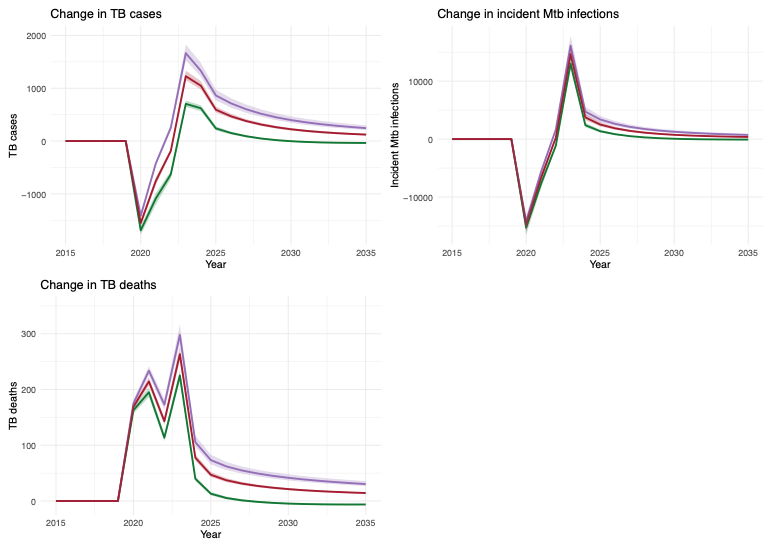

**Figure S2: Model estimated annual difference in TB outcomes under the fitted pandemic scenario compared with each counterfactual immigration scenario during 2015–2035.**

https://www.nature.com/articles/s41467-021-20990-2.pdf.

4.	Centers for Disease Control and Prevention. Available at: http://wonder.cdc.gov/mcd-icd10.html.

5.	United States Centers for Disease Control and Prevention. Online Tuberculosis Information System (OTIS). Available at: https://wonder.cdc.gov/TB-v2022.html. Accessed 23 Apr 2024.

6.	Baugh R. Annual Flow Report U.S. Lawful Permanent Residents: 2022. Department of Homeland Security **2023**.
